# Characterization of a novel, low-cost, scalable ozone gas system for sterilization of N95 respirators and other COVID-19 related use cases.

**DOI:** 10.1101/2020.06.24.20139469

**Authors:** Nikhil Dave, Katie Sue Pascavis, John Patterson, Michael Kozicki, David Wallace, Abhik Chowdhury, Morteza Abbaszadegan, Absar Alum, Pierre Herckes, Zhaobo Zhang, Josh Chang, Clinton Ewell, Tyler Smith, Mark Naufel

## Abstract

Severe acute respiratory syndrome coronavirus 2 (SARS-CoV-2), an elusive and highly pathogenic agent, has resulted in the ongoing COVID-19 pandemic affecting numerous populations worldwide. New studies investigating the tenacity of SARS-CoV-2 have highlighted its ability to persist on a myriad of surfaces for several days, including gowns and shoes. As a result, there is a global need for sterilization of a variety of potentially-contaminated items, ranging from clothing to personal protective equipment like face coverings. To this end, we have designed and constructed a cost-effective, scalable, and sustainable sterilization system that uses ozone gas to inactivate viral particles. We sought to determine the efficacy of the system in the sterilization of viral particles as well as its ability to sterilize N95 respirators for reuse. N95 respirators inoculated with P22 bacteriophage and sterilized in the ozone system showed a 6-log_10_ reduction in viral load when treated at 25 ppm for 150 minutes. Further, N95 respirators treated with five 150-minute cycles at 35 ppm for a total concentration-time product (CT) of 26,250 ppm·min in the ozone system showed comparable filtration efficiency to untreated N95 respirators in a 50 to 200 nmr particulate challenge filtration test. Interestingly, the surgical N95 respirators tested showed complete inactivation of fluid resistance and degradation of the elasticity of polyisoprene straps after five cycles in the sterilization system. Taken together, these data suggest that while our ozone system may negatively affect certain protective aspects of surgical N95 respirators, it does effectively sterilize viral particles and can be utilized for a multitude of other use cases, including sterilizing polypropylene face coverings after potential SARS-CoV-2 contamination. In addition to providing long-term environmental benefits, deployment of this system during the ongoing pandemic reduces the risk of COVID-19 community transmission while conserving monetary resources otherwise spent on the continuous purchase of disposable face coverings.

## Introduction

The COVID-19 epidemic has accumulated well over 4 million cases worldwide to date, and is continuing to grow [1]. Severe acute respiratory syndrome coronavirus 2 (SARS-CoV-2), the pathogen responsible for the COVID-19 epidemic, has been classified as highly contagious through analysis of outbreak dynamics worldwide [2, 3, 4]. Further studies have shown that SARS-CoV-2 has the ability to remain stable on a variety of surfaces, resulting in its ability to spread rapidly [5,6]. Interestingly, recent work has highlighted its ability to remain stable on items such as the shoes of medical care workers, allowing shoes to act as a vector for viral transmission [7]. Given the rapid spread of SARS-CoV-2 and its high stability, there is an increasing need for effective sterilization techniques that can sterilize a myriad of items which may carry SARS-CoV-2. With this specific problem in mind, the terms “sterilization of viral loads” and “sterilization” will be used interchangeably in this work.

The global scientific community has performed extensive work to characterize novel sterilization systems for decontamination of potentially-contaminated items, from disposable N95 respirators to hospital gowns and even cell phones. However, a majority of these techniques are designed and built for high-throughput sterilization in hospital and clinical settings. These systems are often expensive and environmentally unsustainable [8]. To this end, we have designed and developed a novel, low-cost, environmentally sustainable ozone generator which can be easily built into a sterilization system and scaled to a variety of sterilization use cases.

Ozone is a well-characterized biocidal agent, even in relatively low concentrations [9,10]. Ozone may be manufactured by subjecting atmospheric oxygen to high electric fields, which results in ionization and recombination of oxygen radicals due to corona discharge [11]. This process allows ozone to be produced without any chemical feedstock, making it a highly sustainable and scalable disinfectant source. In order to generate the necessary electric field to promote ozone formation, a high-voltage generator and a suitable electrode are required.

The ozone treatment system described in this paper is constructed from a sealed ambient-pressure chamber and a low-cost ozone generator, which when combined, allow a prescribed ozone concentration to be achieved within the chamber for a sustained period of time. The sealed ambient-pressure chamber may be constructed from one of a number of commonly-available plastic tubs or bins. The low-cost ozone generator is constructed from widely-available automotive and consumer electronics, allowing adequate ozone to be generated without specialty components. The ozone treatment system does not require high power input, and may be powered from any 12V DC supply capable of providing at least 2 amps, including automotive power sources in the case where an AC grid supply is not available. In order to ensure a narrow range of ozone levels in the treatment chamber, an ozone concentration sensor may be coupled with the circuitry, but in certain contexts where ozone concentration does not need to be closely controlled, a timer may be used to control the ozone accumulation within the chamber instead.

Table 1 lists the bill of materials required for the construction of one prototype unit of the ozone gas sterilization system. The list of components, quantity of components, and approximate cost (per component in US dollars) of each are tabulated. It should be noted that this list of components does not cover all possible configurations of the system, and gives the required materials for one specific implementation of the system.

**Table 1.** Bill of materials for the construction of one ozone gas sterilization system.

| **Component:** | **Quantity** | **Approximate Cost (Each):** |
| --- | --- | --- |
| Automotive Ignition Coil | 1 | \$3.76 |
| 12V 2A Power Supply | 1 | \$5.50 |
| Silicone Wire (22 AWG) | 2 m | \$1.09 |
| TIP35CW Transistor | 1 | \$0.76 |
| Arduino Nano | 1 | \$3.39 |
| 680uF Capacitors | 2 | \$0.14 |
| LM7805 Linear Regulator | 1 | \$0.50 |
| 120mm DC Fan | 1 | \$6.99 |
| MQ-131 Ozone Sensor | 1 | \$29.99 |
| Zip Ties | 10 | \$0.08 |
| HVAC Aluminum Tape | 0.4 m | \$0.67 |
| Sealed Container | 1 | \$10.99 |
| | **Total Cost:** | **\$65.41** |

In addition to low material cost, the parts used are available outside of medical supply chains. This allows for the system to be scaled to the needs of businesses, educational institutions, and small healthcare providers even during present shortages. With this goal in mind, we pursued validation of the system by quantitatively demonstrating its efficacy for sterilization. Surgical N95 respirators were selected for this study due to their well-defined standards for minimum performance. Additionally, we sought to provide additional validation of this treatment process for use with potential SARS-CoV-2 vector items such as shoes through a qualitative evaluation of degradation under typical treatment conditions.

Validating a system for sterilization of N95 respirators involves not only biological validation but also consideration of the effects of sterilization on NIOSH-dictated standards such as fit, inhalation resistance, and most importantly a N95’s required filtration efficiency of 95% or higher for non-oily solid and liquid aerosols [12,13,14]. Furthermore, any surgical mask such as a surgical N95 must be validated for fluid-resistant performance as regulated by the FDA [12]. To this end, we experimentally validated the system’s virucidal capability, impact on the filtration efficiency of N95 respirators, and effect on the fluid resistance of the N95 respirators.

## Methods

### Validation of ozone concentration produced by generator

A *Thermo 49C* UV photometric ozone analyzer was connected to a sample ozone treatment chamber in order to assess the ozone concentration within the chamber. The rates of ozone production and decay in the test chamber were evaluated using this setup. The ozone generator’s performance was quantified by measuring the maximum ozone generation rate of the ozone generator under continuous operation. This value was determined by measuring the concentration of ozone in this fixed-volume chamber over time. The decay rate of ozone at room temperature was determined by measuring the concentration of ozone in the chamber over time with the ozone generator turned off.

With the MQ-131 ozone concentration sensor connected to the microcontroller, an ozone setpoint of 25 ppm was set in the microcontroller programming after initial calibration against a metrology-grade ozone analyzer. The ozone concentration was then measured over the entire duration of a 150-minute typical treatment cycle. It was determined in this step that the sensitivity of the MQ-131 ozone sensor was subject to change over time, so a calibration curve of the chamber concentration over time was plotted. In a future implementation of the microcontroller program, this calibration curve may be incorporated to ensure that the ozone level remains close to the setpoint within the chamber.

### Validation of virucidal activity using P22 bacteriophage as a proxy for SARS-CoV-2 Viral Preparation and Assay

Bacteriophage P22 (ATCC® 19585-B1™) was propagated using *Salmonella typhimurium* (ATCC® 19585™) as the host bacterium using the double agar layer (DAL) technique. Briefly, 1 mL of the sample and 1 mL of the host cell bacteria, in the log-phase of growth, were added to a 5 mL of melted top agar in a test tube which was kept in a water bath at 48°C. The mixture was gently poured onto a bottom agar plate and kept undisturbed to let the top agar to solidify. Then, the plate was incubated upside down at 37°C and plaques were counted after 24 hours of incubation. A positive and a negative control was included in every DAL assay.

The bottom agar plates were prepared using Tryptic Soy Agar (TSA) (Difco Laboratories, Division of Becton Dickinson & Co.). Prepared TSA was sterilized using an autoclave, cooled to 55°C and then dispensed into petri plates (20 ml per plate). Plated TSA was allowed to solidify and then stored at 4°C until used.

The top agar was prepared using Tryptic Soy Broth (TSB) (Difco) by adding 0.7% agar (Difco). Five mL of the top agar medium were dispensed to test tubes, capped and then autoclaved. The top agar tubes were stored at 4°C until used.

### Inoculation and Test Procedure

A surgical 3M 1860 N95 mask was cut to 2.5 x 2.5 cm pieces using a sterilized pair of scissors to obtain test coupons. Three coupons were placed in a sterilized petri dish, and each was inoculated with P22 bacteriophages at a total concentration of 1.6×10^8^ PFUs per coupon by transferring 100 µL of the stock solution on their inner surface. The inner surface of the mask was selected based on its higher absorbance capability compared to the outer surface which is highly hydrophobic (data not shown). The inoculated coupons were allowed to dry at room temperature for 5 minutes to equilibrate with the mask material.

Triplicate sets of inoculated coupons were placed in the ozone treatment system. Both systems were operated according to the parameters specified earlier.

After the operation cycle completed, test coupons were retrieved from the systems. Each coupon was placed in a 50 mL tube containing 30 mL of elution buffer. The tube was vortexed to recover viruses from the coupon, and then the recovered buffer was analyzed using DAL technique.

### Validation of N95 respirator filtration efficiency before and after treatment

The capture efficiency tests were performed using a custom set-up. Challenge aerosols were generated using a medical nebulizer (drive medical, Port Washington, NY, USA) from aqueous solutions. Tests were performed using fumed silica nanoparticle slurry solutions which have been extensively characterized [15]. The nebulization resulted in a broad challenge aerosol distribution and typical observational range was from 50-200 nm, a range which incorporates particles smaller than individual virions. This challenge aerosol covers the range of aerosols used in the NIOSH tests methods, 75 ± 20 nm NaCl particles for N type masks [16] and dioctyl phthalate (185 ± 20 nm) particles for P99 masks [17]. The challenge aerosol was passed through a trap bottle to remove larger particles. The aerosol then was measured directly or passed through a 25 mm diameter punch sample of the mask material, held in a filter cassette. The size resolved particle number concentration were determined with a Scanning Mobility Particle Sizer (SMPS) set-up consisting of a TSI 3088 Soft X-Ray neutralizer, a TSI 3082 aerosol classifier, a TSI 3085A Nano differential mobility analyzer (DMA) and a TSI 3752 high concentration condensation particle counter (CPC) (TSI, Shoreview, MN, USA). All tests were run in triplicate and for at least 10 minutes each. The respirators used for validation were surgical 3M 1860 N95s.

### Validation of N95 respirator fluid resistance before and after treatment

The fluid resistance of the N95 mask surface of control-group (untreated) and experimental-group (5 cycles of ozone treatment) N95 masks was quantified by means of a water drop deflection test. Distilled water was dropped from a fixed height of 17 cm above the base of the mask onto the surface of a mask. The mask was tilted such that the nose-clip edge of the mask was elevated 5 cm above the base of the mask, as in Fig. 1. It was also qualitatively noted whether any absorption of the water drops into the mask surface was observable. While the validation process was not as intensive as to meet the ASTM Test Method F1862 “Resistance of Medical Face Masks to Penetration by Synthetic Blood” as required by the FDA to deem a mask as “surgical,” both procedures utilize visible penetration of liquid to determine the effectiveness of the fluid resistant layer [12]. The respirators used for validation were surgical 3M 1860 N95s.

**Fig. 1.**
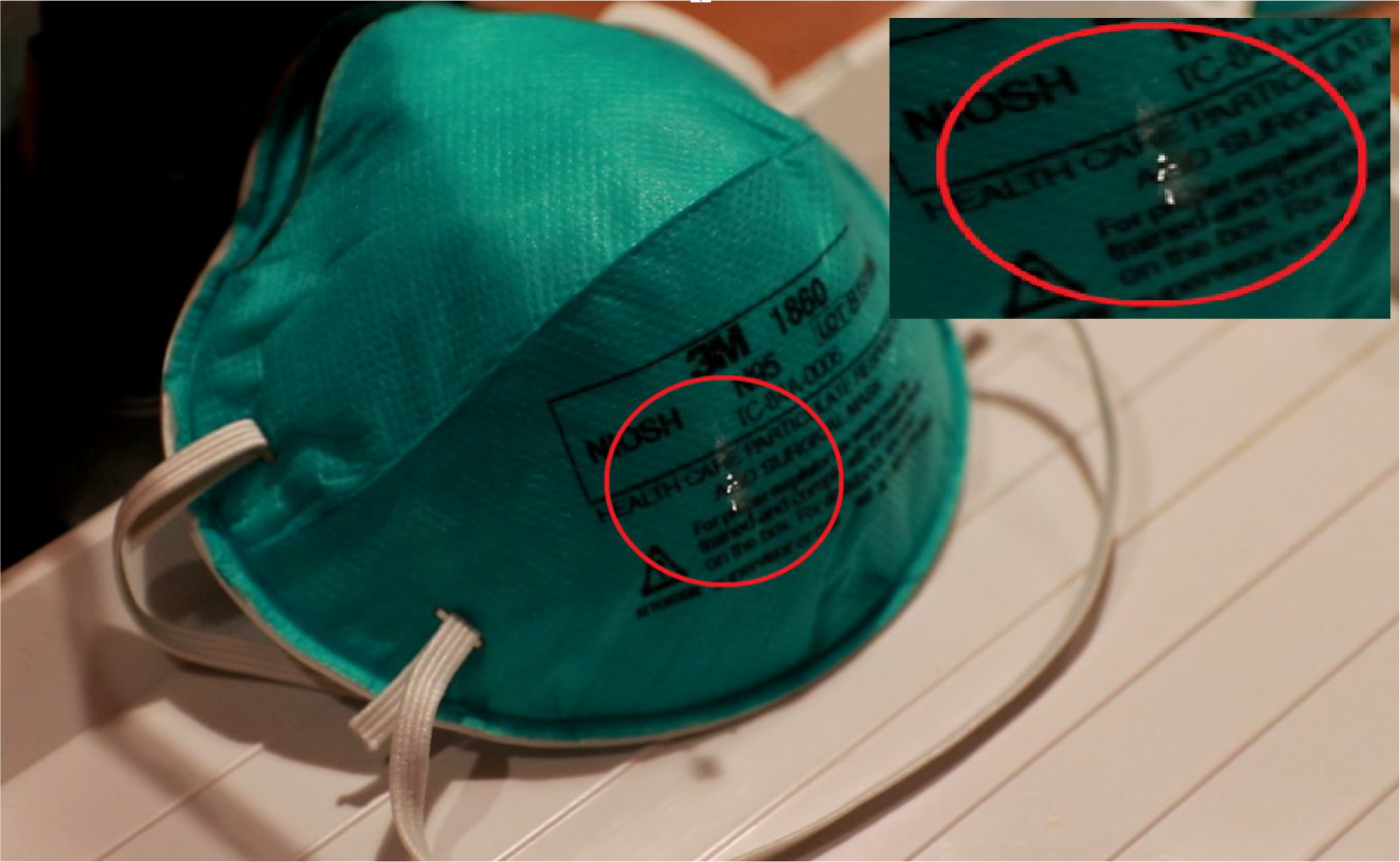
The fluid resistance validation test setup, showing the water drop about to encounter the hydrophobic layer of the mask surface.

## Results

### Validation of ozone concentration produced by generator

Initial testing with a prototype of the ozone generator yielded a production rate of 150 mg/hr, and ozone concentrations within the test chamber exceeding 150 ppm were achieved. A plot of ozone generation is shown in Fig. 2 and a plot of ozone decay is shown in Fig. 3 for the prototype ozone generator in a 4-cubic foot container as measured using the *Thermo 49C*.

**Fig. 2.**
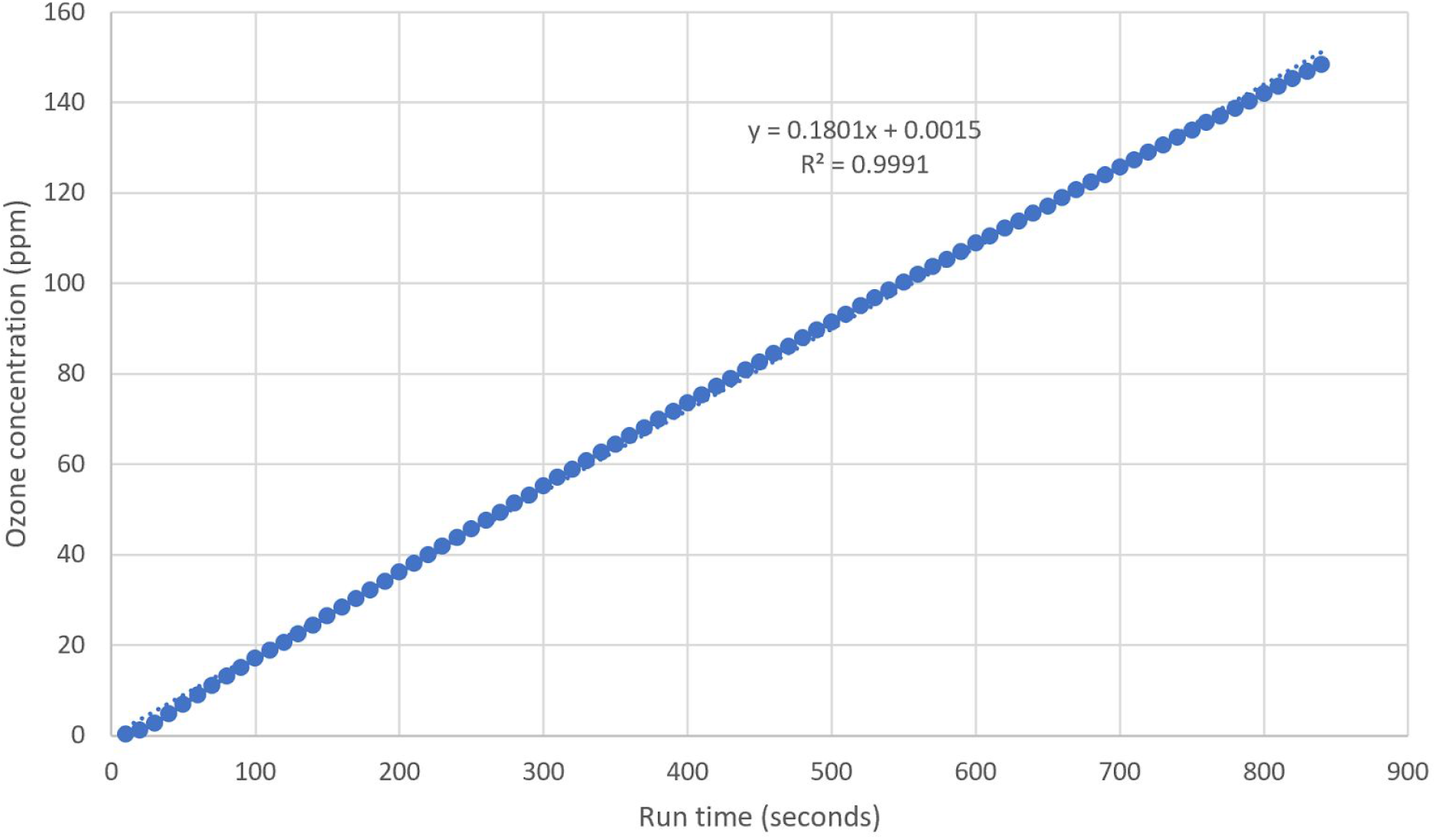
Plot of ozone concentration over time in a 4-cubic foot chamber with ozone generator continuously operating.

**Fig. 3.**
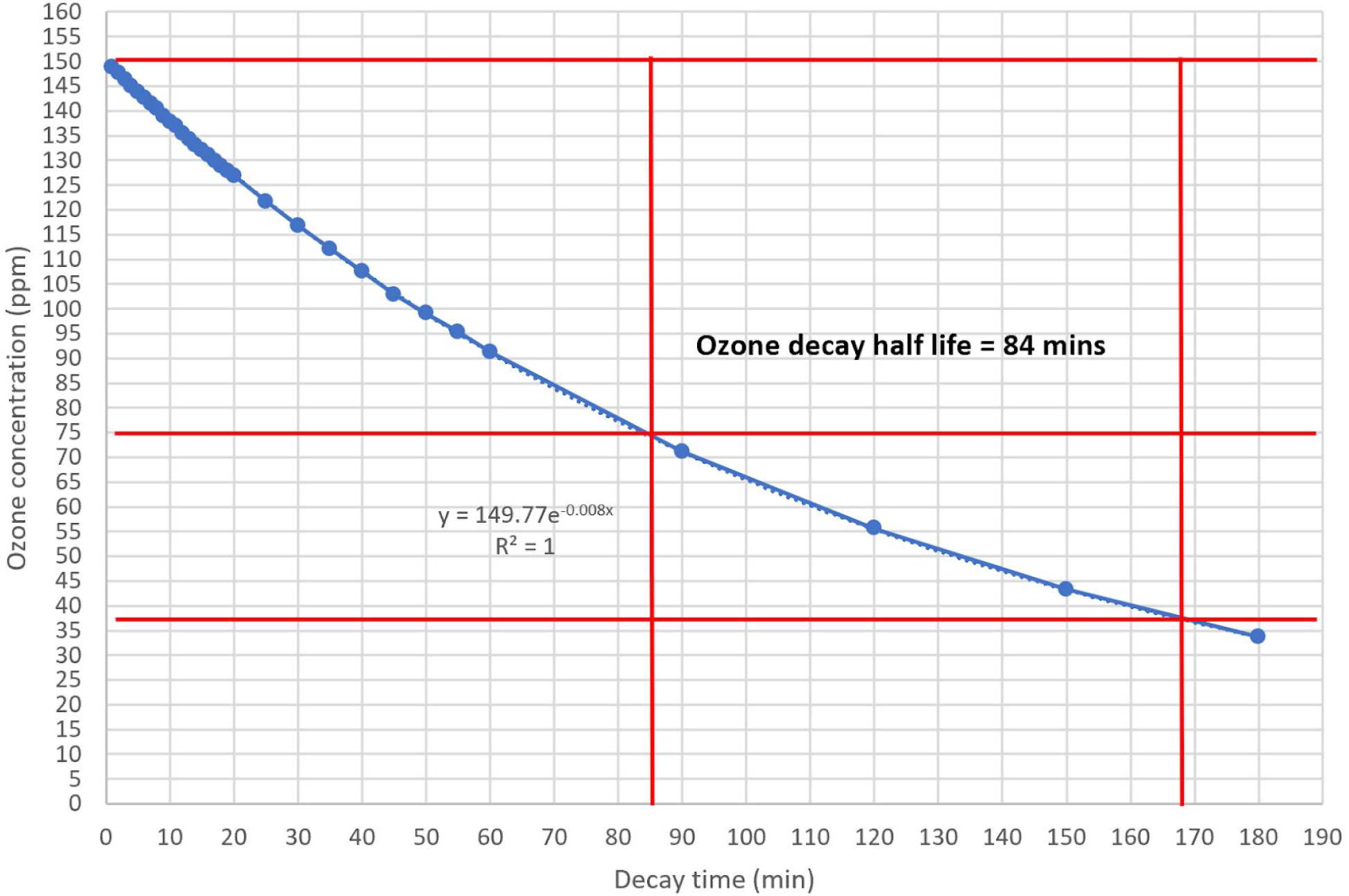
Plot of ozone concentration over time in a 4-cubic foot chamber with ozone generator turned off (decay mode). Red lines placed to assist in calculation of decay time.

Target concentrations for treatment range from 20 to 30 ppm. These results indicate that the ozone generator will provide adequate ozone to the chamber to maintain this concentration range throughout the duration of the treatment cycle.

It is important to note that the material composition of items placed into the ozone treatment chamber may have a profound effect on the decay rate of the ozone concentration. Therefore, the addition of the MQ-131 ozone sensor to the system is desirable in order to control the ozone concentration within the chamber in a specified range, regardless of the ozone consumption rate of the items in the chamber.

The overall system, including the *Sterilite* chamber, the complete ozone generator system, and the MQ-131 ozone sensor, were operated for a typical treatment cycle period of 150 minutes. The ozone concentration within the chamber was measured every 10 minutes using the *Thermo 49C*, and the resulting concentration curve is shown in Fig. 4.

**Fig. 4.**
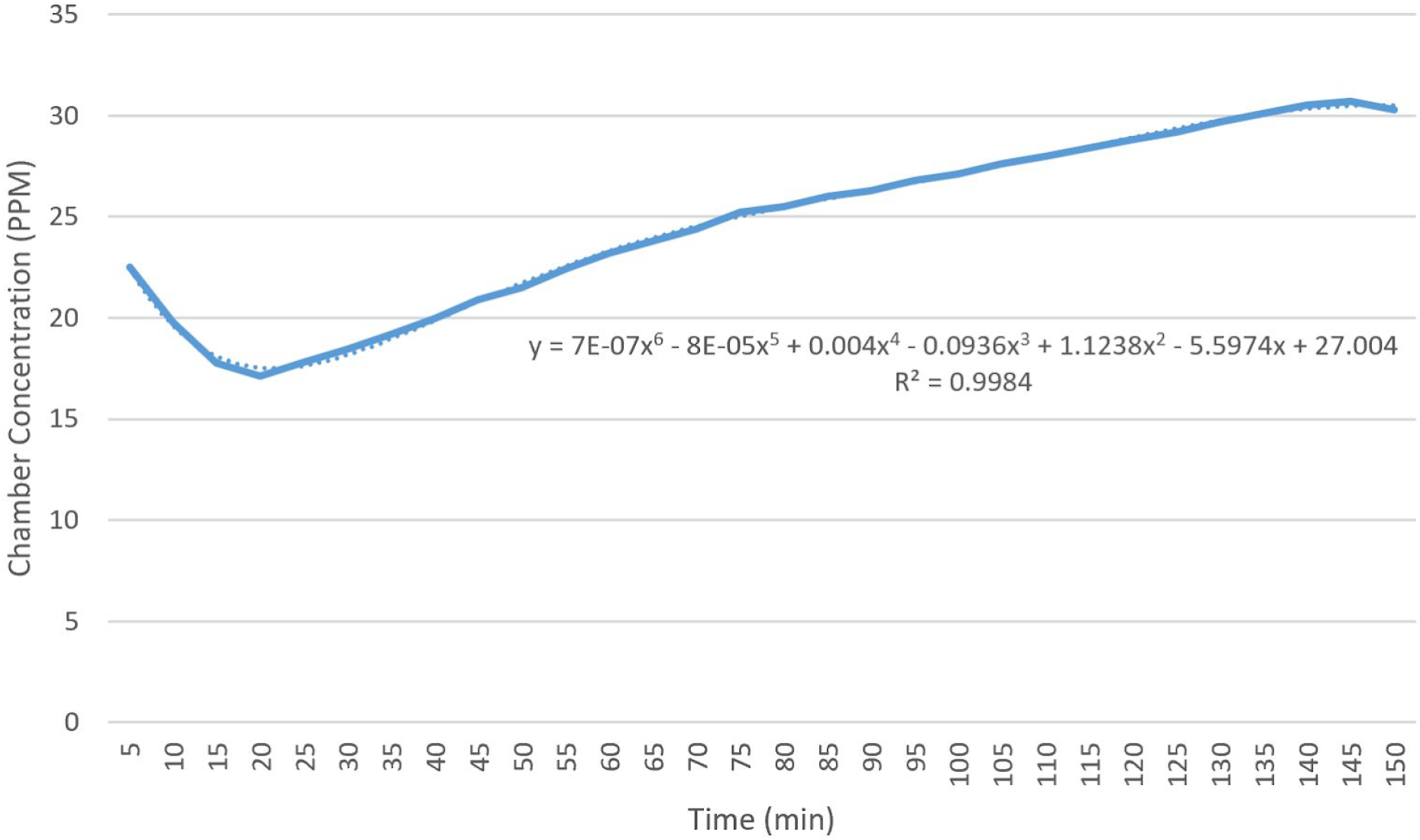
Plot of ozone concentration over time under closed-loop control using the MQ-131 sensor and a setpoint of 25 ppm.

It is evident from the concentration curve of Fig. 4 that the sensitivity of the MQ-131 ozone sensor drifts over time. Because the average chamber concentration is approximately equal to the set point of 25 ppm, the chamber may optionally be operated without software calibration. The chamber calibration curve may vary from one test run to the next, so calibration of the sensor using a precision ozone measurement system such as a *Thermo 49C* ozone analyzer is recommended if high ozone concentration precision is required.

### Validation of virucidal activity using P22 bacteriophages as a proxy for SARS-CoV-2

The viral inactivation data is presented in Table 2. The ozone system achieves sterilization of viruses on N95 mask coupons. In all tests, greater than 6-log_10_ reductions of P22 bacteriophage were achieved under the test conditions, which surpasses requirements for sterilization [18, 19]. The test coupons of inoculated N95 respirator material were exposed to an average ozone concentration of 25 ppm for 150 minutes, resulting in a total dose of 3750 ppm·min being administered to these samples. The observed 6-log_10_ reduction of the viral proxy–a similar result to Yale University’s validation of the BQ-50 VHP system (Bioquell, Horsham, PA) and NIOSH and CDC’s evaluation of UVGI for sterilization of N95s–demonstrates the system’s effectiveness against SARS-CoV-2 [12, 19].

**Table 2.** Inactivation of P22 bacteriophages on N95 mask coupons using ozone or vaporized hydrogen peroxide.

| **Treatment** | **Replicate** | **Initial**<br>**$\log_{10}$ Inoculated** | **Final**<br>**$\log_{10}$ Recovered** | **$\log_{10}$**<br>**Reduction** |
| --- | --- | --- | --- | --- |
| **Ozone** | 1 | 8.20 | 1.78 | 6.43 |
|  | 2 | 8.20 | 1.78 | 6.43 |
|  | 3 | 8.20 | 1.48 | 6.73 |

### Validation of N95 respirator filtration efficiency before and after treatment

The filtration validation study described in the methods section indicated that at the challenge aerosol sizes tested, the measured capture efficiency of one sample respirator treated with 5 ozone treatment cycles under the procedures recommended herein remained above the minimum filtration efficiency value of 95% required for the respirators to be classified as N95 type (the NIOSH validation test uses similar particles but different operating conditions) [14]. The worst-case value of measured filtration efficiency for a 5-cycle treated N95 respirator was found to be 98%. These results indicate a minor reduction in filtration efficiency at lower particulate sizes after several cycles through the recommended ozone treatment process, but they are indicative that the minimum filtration efficiency required for N95 respirator classification is most likely to be preserved over a wide range of particulate sizes even beyond five treatment cycles. The results of this filtration efficiency testing are depicted in Fig. 5.

**Fig. 5.**
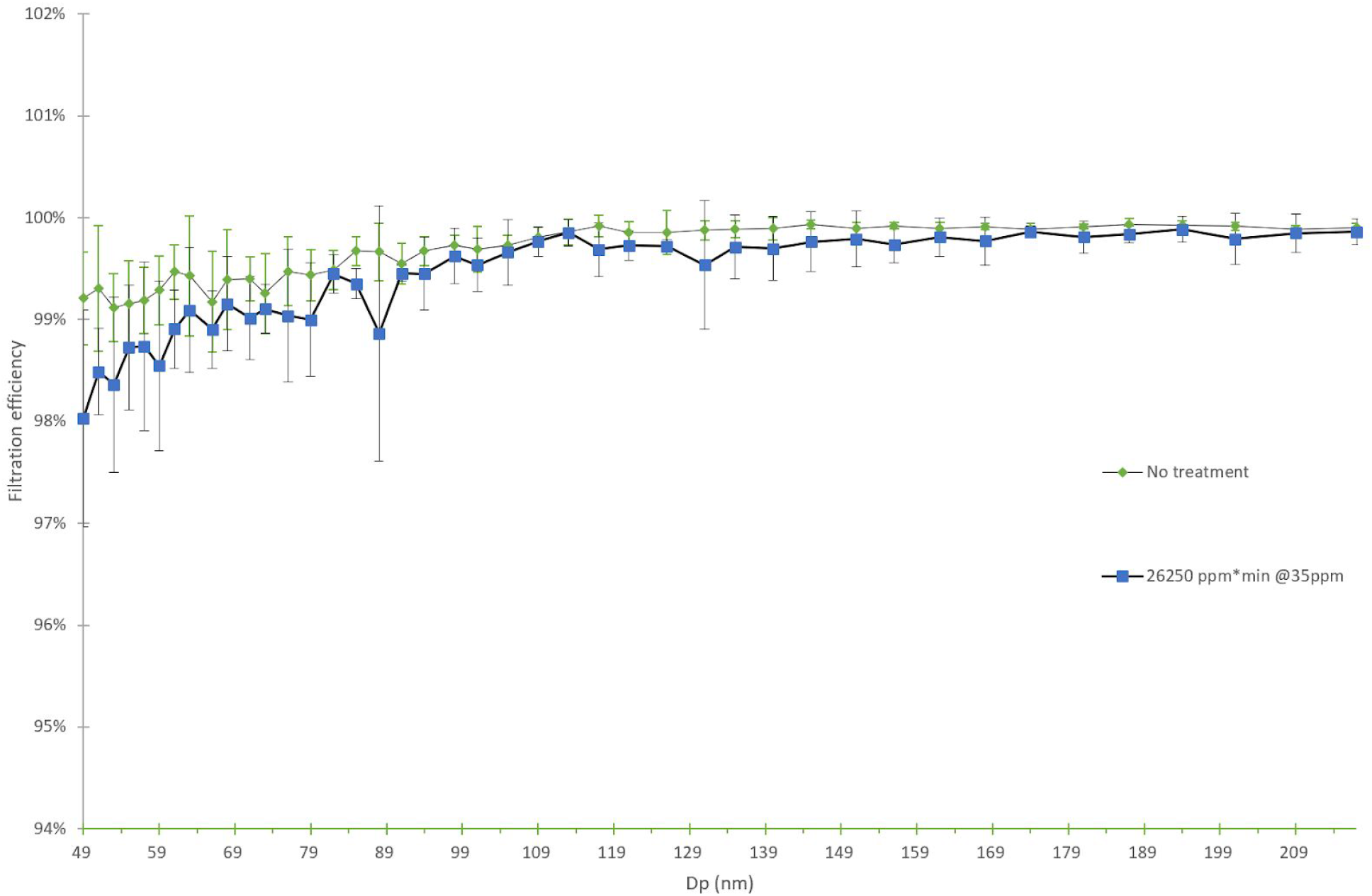
Filtration efficiency of untreated N95 respirators, N95 respirators treated with 26250 ppm·min ozone exposure from 50 nanometers to 300 nanometers.

### Validation of N95 respirator fluid resistance before and after treatment

Initial testing of N95 surgical masks that have been treated with three and five cycles of ozone treatment showed severe degradation of the hydrophobic layer on the surface of the mask. A drop deflection test was not possible, as droplets of water quickly absorbed into the mask surface in the samples tested. Therefore, it has been shown that the hydrophobic layer of some surgical N95 masks is destroyed by the ozone treatment process.

## Discussion

Sterilization of healthcare-related items is commonly achieved through the use of gas-phase disinfectants such as ethylene oxide (EtO), nitrogen dioxide, peracetic acid, ortho-phthalaldehyde (OPA), and glutaraldehyde (Cidex) [21]. To this end, many common methods used for decontamination of a variety of items in hospitals and other sterile environments require an existing feedstock of a sterilization agent. In contrast, our sterilization system requires no existing feedstock to produce the sterilization agent and only needs a power supply to operate continuously. This generates significantly fewer waste materials produced in comparison to orthodox clinical high-throughput sterilization procedures. Similarly, using the ozone system over common clinical decontamination methods also significantly reduces the resource costs associated with decontamination and sterilization. Moreover, many of the aforementioned sterilization agents require a relatively long treatment time. For example, commonly used gas-phase EtO requires a treatment time of over 12 hours [21]. In contrast, this ozone system was shown to sterilize P22-inoculated N95 respirators in 1 hour, suggesting that the system is also more time efficient in sterilization. As such, we believe that the ozone system described in this paper will provide first responders, businesses, educational institutions, and healthcare facilities with unprecedented access to easy-to-use, affordable sterilization technology.

To verify the performance of this device, the system’s virucidal efficacy, impact on the filtration efficiency of N95 respirators, and compatibility with fluid-resistant N95 respirators were analyzed experimentally. Measurements taken from a sample ozone treatment chamber using a *Thermo 49* ozone analyzer indicated that under closed-loop control for one treatment cycle, a concentration of ozone ranging from 20 – 32 ppm is maintained with the proposed system design after an initial warm-up period of 45 minutes. In a biological validation test in which P22 bacteriophage was utilized as a surrogate for SARS-CoV-2, an exposure time of 150 minutes at 25±5 ppm ozone concentration was shown to provide greater than 6-log_10_ reduction in viral load, similar to Yale University’s methodology for validating the Bioquell sterilization system for use during the COVID-19 epidemic [20]. This result not only meets but exceeds the ASTM definition for sterilization [19].

Furthermore, N95 respirators treated with 5 cycles of 150-minute exposure at 25±5 ppm ozone concentration remained above the minimum required filtration efficiency of 95% for this type of respirator, indicating that ozone does not pose a significant risk to filtration efficiency in the concentrations, time, and number of cycles studied.

Conversely, the ozone CT tested did have a profound effect on the outer hydrophobic coating of surgical N95 respirators, which showed substantial loss of fluid resistance in a water droplet test after less than 3 cycles of ozone treatment. Similarly, polyisoprene elastic bands showed visible degradation after exposure to multiple sterilization cycles. While these negative effects can be circumvented by applying a replacement hydrophobic layer, (e.g., Scotchgard™ Fabric Protector, 3M Company-Cat. No. 4101, 4106) after each sterilization cycle and coating elastic straps in a ozone-resistant substance during treatment (e.g., mineral oil or petrolatum), the ozone sterilization system is not recommended for the sterilization of this type of surgical N95 respirator without further testing. Other types of N95 respirator may show greater degrees of compatibility with this process; further research into compatible options should be pursued.

While further research into the compatibility of ozone and hydrophobic materials is conducted, the sterilization and filtration results suggest that this system is well-suited for processing most face coverings currently available to the general public. Additionally, the system may be suitable for use with many hard materials typically found in household and commercial settings, such as tools, office supplies, and other frequently-handled items. In particular, items consisting of stainless steel, aluminum, PVC, nylon, teflon, silicone, plexiglass, borosilicate glass have been shown to be compatible with ozone sterilization processes [21]. Rubber, latex, brass, zinc, and nickel-based materials are not recommended for use in this ozone sterilization system [21].

Finally, the real-world limitations of ozone sterilization for personalized protective equipment (PPE) and other items must be considered. While shoes and other items demonstrated no visible degradation after sterilization (see Supporting Information), outside wear and contamination could decrease the durability of ozone-treated items. Precautions against wear of use, such as examining sterilized articles for deformation or sterilizing for less than the validated number of cycles, should be taken.

Another consideration for real-world use is the potential impact of ozone gas on human health. For safety, OSHA requires ozone gas exposure to remain below .1 ppm over an 8-hour, time-weighted average [22]. We have shown that in a typical occupied space with limited ventilation, ozone concentrations 15 minutes after each treatment cycle remain much lower than maximum the OSHA permissible exposure limit (PEL), indicating the safety of the described system (see Supporting Information) [22]. Nevertheless, after a treatment cycle, one should turn their head away from the system when opening the chamber, allow the gas to disperse for 10 minutes before removing any items, and only open the system in a well-ventilated space [23].

## Conclusions

Achieving a 6-log_10_ reduction in P22 bacteriophage viral loads suggests that our ozone system will be effective in eliminating SARS-CoV-2 on various items including PPE. Furthermore, decontaminated N95 respirators still consistently exceeded standard filtration performance as required by NIOSH standards. While the degradation of the elastic straps and fluid resistant layer of N95 respirators requires additional considerations before N95 respirators may be safely reused in certain environments, the 6-log_10_ reduction of viral loads and minimal effect on filtration performance demonstrates that our ozone system can serve as a cost-effective sterilization tool for certain applications. Such applications include the sterilization of non-hydrophobic masks or other face coverings, as well as potentially-contaminated items worn in medical environments such as hospital gowns and shoes. Moreover, given the efficacy of the tested system against P22 bacteriophage loads, ozone can be used to sterilize other compatible items outside of the clinic. For example, businesses operating during the COVID-19 pandemic can use our ozone sterilization system to sterilize face coverings for reuse, thus reducing the financial burden of purchasing new face coverings and inspiring both employee and consumer confidence. With growing demand for surgical masks and other face coverings due to community use, widespread ozone sterilization can also serve as a tool to reduce the impact of this newfound demand on the personal protective equipment supply chain.

Beyond the COVID-19 epidemic, our novel ozone generator offers a cost-effective solution to the environmental toll endured from the widespread use of waste-generating decontamination agents. Present decontamination agents present significant monetary costs and result in the generation of large quantities of biohazard and chemical waste. In contrast, our ozone system requires no initial chemical feedstock and does not generate any significant biohazard waste. Furthermore, the system’s lack of feedstock allows it to be used wherever power is available. This, combined with the system’s use of affordable parts from outside medical supply chains, makes it a scalable solution even during the ongoing pandemic. Large-scale adoption of our ozone system, where appropriate, can allow users to effectively decontaminate materials while also conserving capital dedicated to sterilization supplies and reducing the environmental footprint left behind from high-throughput decontamination.

## Data Availability

All raw data is available upon request.

## Acknowledgements

Capture efficiency testing was supported by the NSF RAPID program (CBET-2028074). Other funding support was provided by the Arizona State University Knowledge Enterprise and the NSF Water and Environmental Technology (WET) Center at Arizona State University (award number 1361815).

## Supporting Information

### Ozone release testing in typical occupied space

In order to establish the effect of operating the ozone sterilization system in a room with limited ventilation, an experiment was carried out in which the ozone sterilization system was placed into a room with no direct ventilation alongside a *Thermo 49C* ozone measuring system. The test was intended to determine if, under typical worst-case usage conditions, the operation of the ozone sterilization system raises ambient concentrations of ozone in an occupied space to levels exceeding the OSHA permissible exposure limit (PEL) of 300 parts per billion (PPB) over a 15-minute window [24].

Testing was performed in a 2.7-meter by 2.1-meter room with a 0.61-meter by 0.61-meter cabinet in one corner. The ceiling height of the room was 2.4 meters. The approximate total volume of the room was 12.7 cubic meters. Special care was taken to ensure that the building HVAC system was disabled throughout each period of testing, and that all exhaust fans in the area were turned off, windows were closed, and no sources of draft were present. The *Thermo 49C* intake hose was positioned near the center of the room approximately 1 meter from the floor during room sampling.

The *Thermo 49C* ozone analyzer was first connected to power and allowed to warm up. A background level of ozone was measured in the room prior to each testing round. The ozone sterilization system was then powered and allowed to operate until the *Thermo 49C* ozone analyzer indicated it had reached an internal ozone concentration of 30,000 PPB (30 ppm) as measured from the sampling port on the treatment chamber. The intake of the *Thermo 49C* was then moved to the room air sampling location, the power was disconnected from the ozone generator, and the lid was removed. The experimenter promptly left the room and closed the door, allowing the room to stand for 15 minutes to allow ozone gas to diffuse evenly throughout the room. After 15 minutes, the door was opened and the most recent ozone concentration measured by the *Thermo 49C* ozone analyzer was recorded.

This experiment was repeated three times. The building HVAC system was operated after each test to remove most of the ozone from the room. To account for changes in the background level of ozone in the testing room due to previous tests, the difference between the initial and final ozone levels in each test was tabulated. The results of each of these tests can be found in Table 3.

**Table 3.**
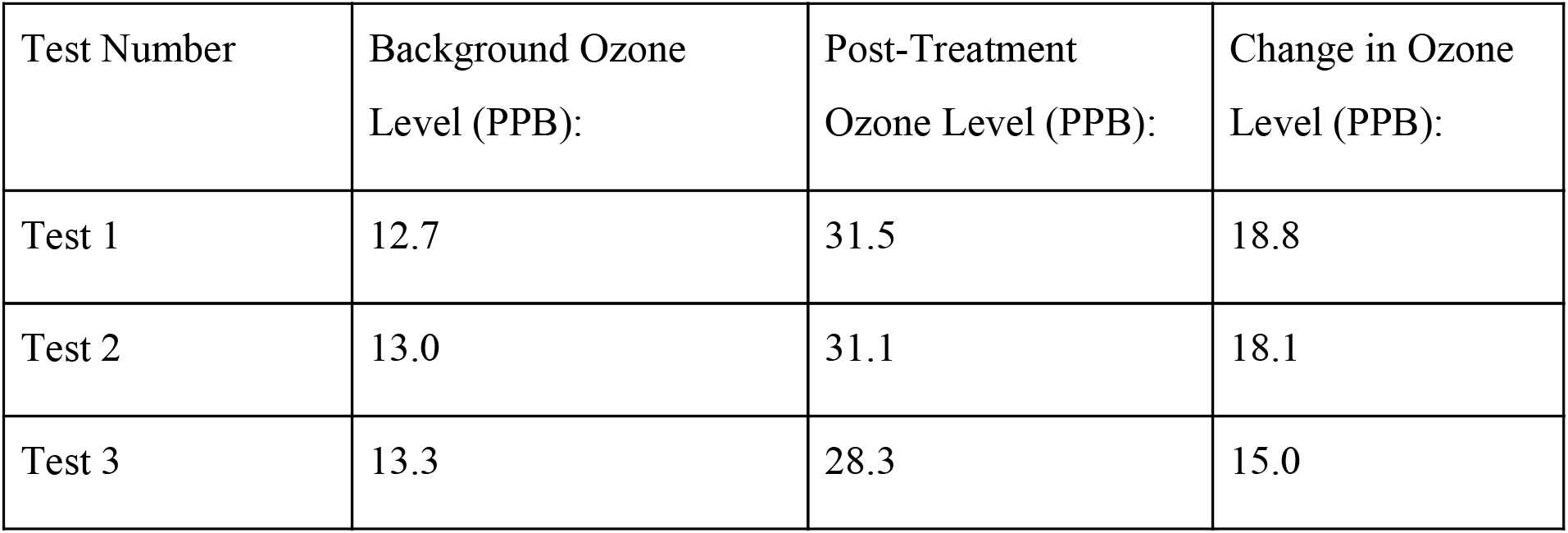
20-minute room ozone concentration after each ozone release test.

| Test Number | Background Ozone Level (PPB): | Post-Treatment Ozone Level (PPB): | Change in Ozone Level (PPB): |
| --- | --- | --- | --- |
| Test 1 | 12.7 | 31.5 | 18.8 |
| Test 2 | 13.0 | 31.1 | 18.1 |
| Test 3 | 13.3 | 28.3 | 15.0 |

**Table 4.** Dosage chart displaying recommended treatment times at 20 ppm of ozone gas to achieve specific reductions in viral load.

| Level of Inactivation of Viral Load Achieved | Treatment Length (min)<br>@ 20ppm of Ozone Gas | Dosage Achieved<br>(ppm · min) |
| --- | --- | --- |
| 90% | 17.5 | 350 |
| 99% | 35 | 700 |
| 99.9%* | 52.5 | 1,050 |
| 99.99% | 90 | 1,800 |
| 99.999% ** | 135 | 2,700 |
| >99.999% | 150 | 3,000 |

\* 99.9% inactivation, also known as a 4- $\log_{10}$ reduction, achieves sanitization of the viral load.
\*\* 99.999% inactivation, or a 6- $\log_{10}$ reduction, achieves sterilization of the viral load, which would be the required level of disinfection for an N95 or equivalent respirator, especially in a healthcare setting [21].

Because the ozone level within the test room never exceeded the OSHA PEL of 300 PPB over a 15-minute window (and remained approximately 10 times lower than this limit), it is likely that the operation of the system is safe indoors under typical operating conditions. It is still recommended that the system be operated in a well-ventilated area, and that the user avoid directly inhaling the exhaust gases released during opening of the container.

### Supplementary qualitative materials testing methods

In order to assess the effects of ozone exposure on certain types of shoe, fabric material, and shoe covering, an ozonation chamber was prepared using one of the Luminosity Lab low-cost modular ozone generator units. A 22.7-liter cooler was used as the sealed chamber, and ozone concentration was determined by interpolation of prior characterization curves.

The ozone generator was initially configured for a 16.6% duty cycle, allowing 10 seconds of ozone generation time for every 50 seconds of ozone decay time. This value was selected before the decay dynamics of the ozone chamber were well-understood, and for the purposes of this report may be considered arbitrary. This duty cycle preserves the approximately linear ozone concentration rise, as it is still within an order of magnitude of the continuous-operation regime. The shoes and material samples to be tested included the following:

1 US Polo Assn “Converse”-style shoe, model number 21629601A

1 Adidas “Ultra Boost” running shoe, model number 606001

1 Saucony “Cohesion 10” running shoe, model number S25354-1

1 Reef Flip-Flop, unknown composition, no model number available

1 confirmed EVA foam flip, no model number available

2 elastic shoe coverings, different manufacturers

1 cloth sample, 100% nylon on one side and 100% polyester on the other side 1 latex balloon

The test chamber was operated for 1 hour per test cycle to allow sufficient time for ozone concentration to rise and ozone to diffuse into the materials under test. The final ozone concentration can be computed according to the previous linear approximation as follows:

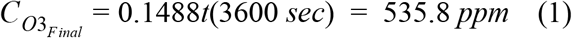

Under the linear approximation of ozone concentration rise, the average ozone concentration throughout the treatment process can be computed as follows:

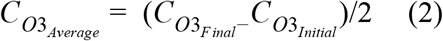

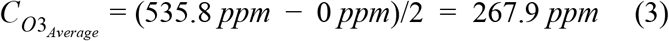

The total approximate dose (in ppm·min) to which the test items were exposed is therefore:

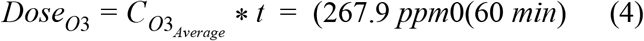

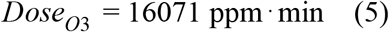

### Supplementary qualitative materials testing observations

Both visual inspection and functionality inspection did not show any signs of degradation on any of the shoe samples tested after exposure to 16071 ppm·min of ozone. This is indicative that the materials from which these shoes are composed are likely not heavily affected by ozone in the dosages that would be commonly applied for sterilization purposes.

The elastic shoe cover from one of the manufacturers (darker blue) showed marked signs of degradation of the elastic after the 16071 ppm·min ozone exposure test. The elastic shoe cover from the other manufacturer did not show evidence of elastic degradation during the treatment period. The non-elastic fabric material did not show signs of degradation.

The nylon and polyester fabric materials did not show signs of degradation after the 16071 ppm·min ozone exposure.

The latex balloon showed extreme deterioration after the 16071 ppm·min ozone exposure. A crack formed in the material and was easily expanded by stretching. The latex took on a sticky consistency and a strong burnt odor.

### System construction

The ozone treatment system may be constructed by assembling the ozone generator system on the lid of the sealed container as shown in Fig. 6. The base of the sealed container may be left unmodified. All electronics may be mounted to the lid of the container by drilling two holes in the lid spaced on either side of each component, and running one or more zip ties through the holes and around the electronic components.

**Fig. 6.**
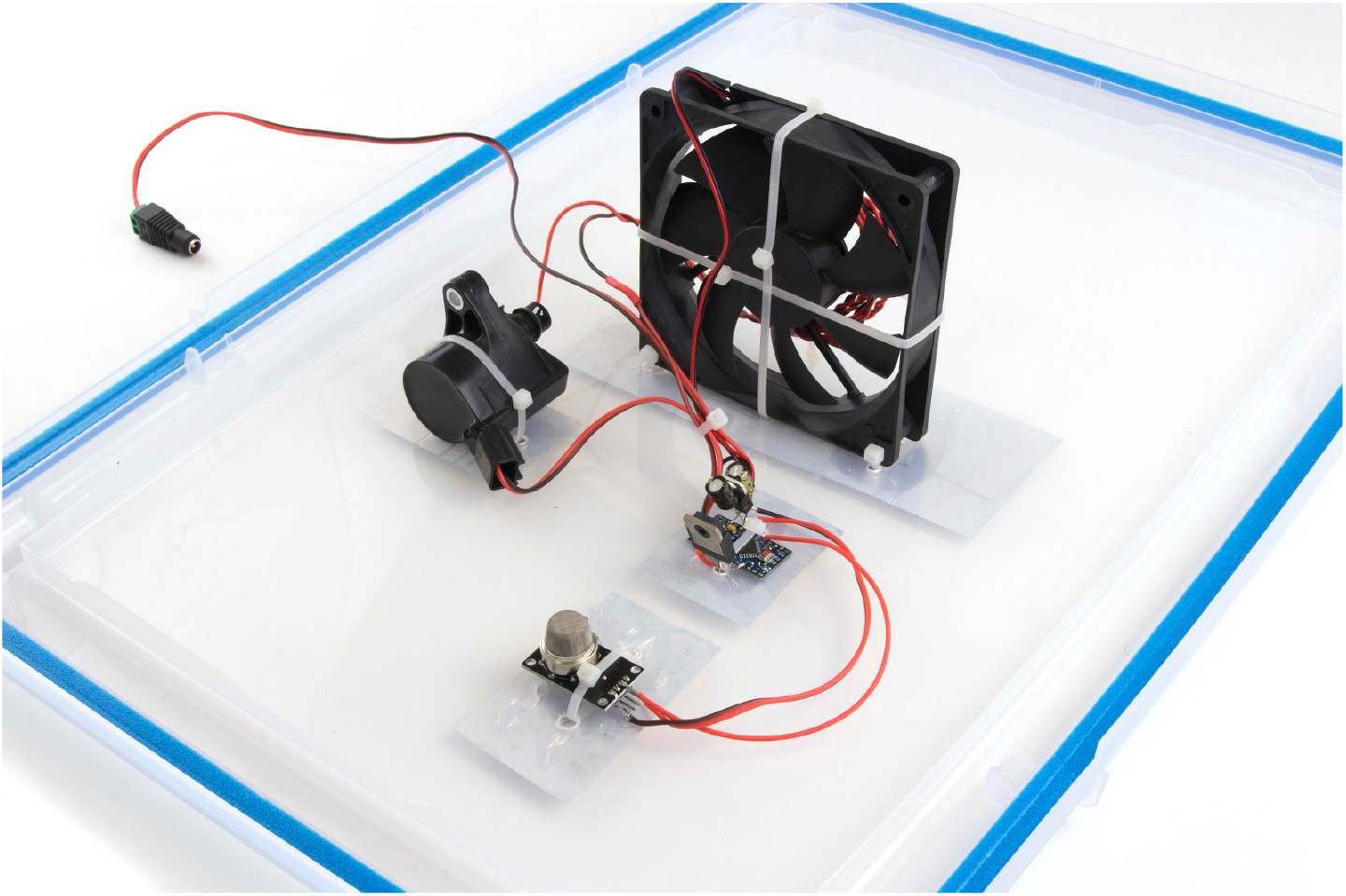
Depiction of the layout of electronics on the inside of a sample ozone gas sterilization system.

Before mounting the components of the ozone generator to the lid of the sealed container, the electronics should be soldered together and assembled as per the wiring diagram depicted in Fig. 7. The silicone wire coil should be affixed to the fan by two crossed zip ties, as shown in Fig. 8. The coil in Fig. 8 is shown operating under long-exposure photographic conditions, but it is not necessary to see this degree of discharge during normal operation. Use approximately 1.5 meters of each conductor in the silicone wire electrode, and twist them together so as to provide approximately 0.5-1mm separation between the wires at the widest separation points. The exact spacing of the silicone wires is not critical, as the closed-loop control will ensure a stable ozone level regardless of the exact generation rate. Ensure that the high-voltage electrode conductors do not directly contact any of the low-voltage electronics or their wiring connections, such as the MQ-131, Arduino Nano, or DC power source. Optionally, a hole may be drilled to allow a DC power jack to be mounted in the lid of the container.

**Fig. 7.**
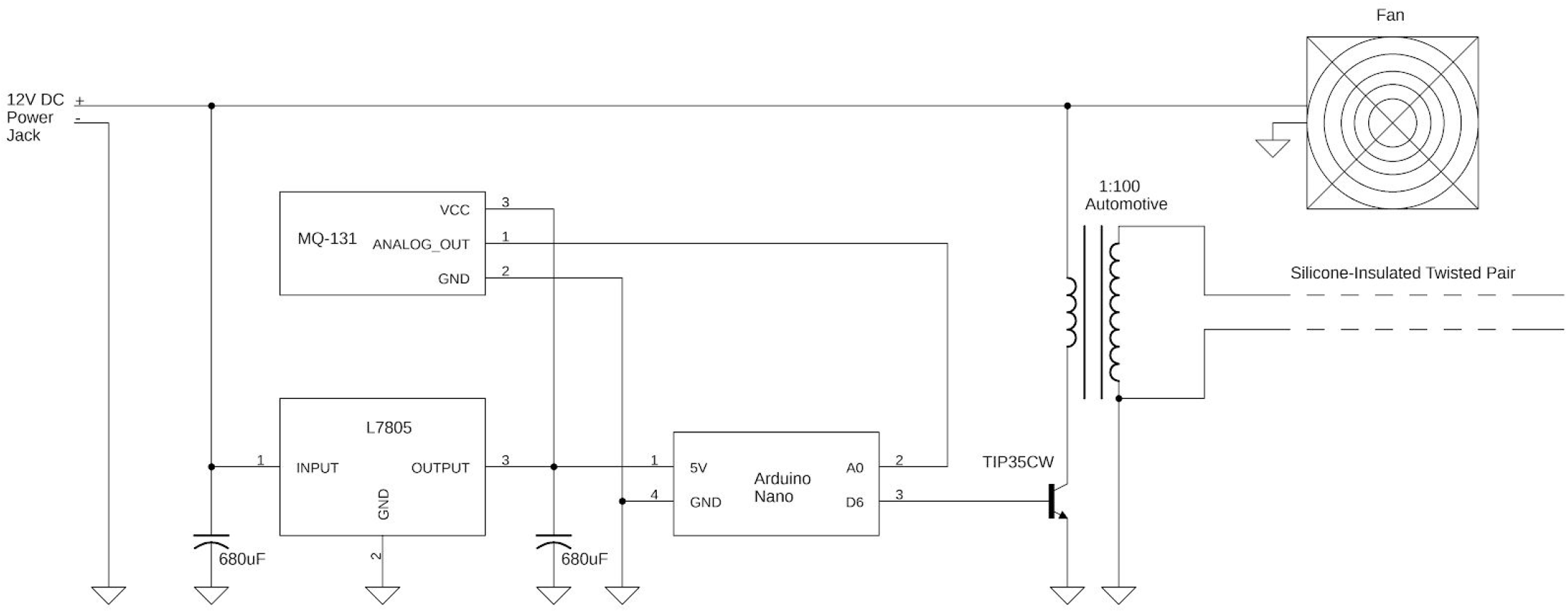
Wiring diagram of the closed-loop implementation of the ozone gas sterilization system.

**Fig. 8.**
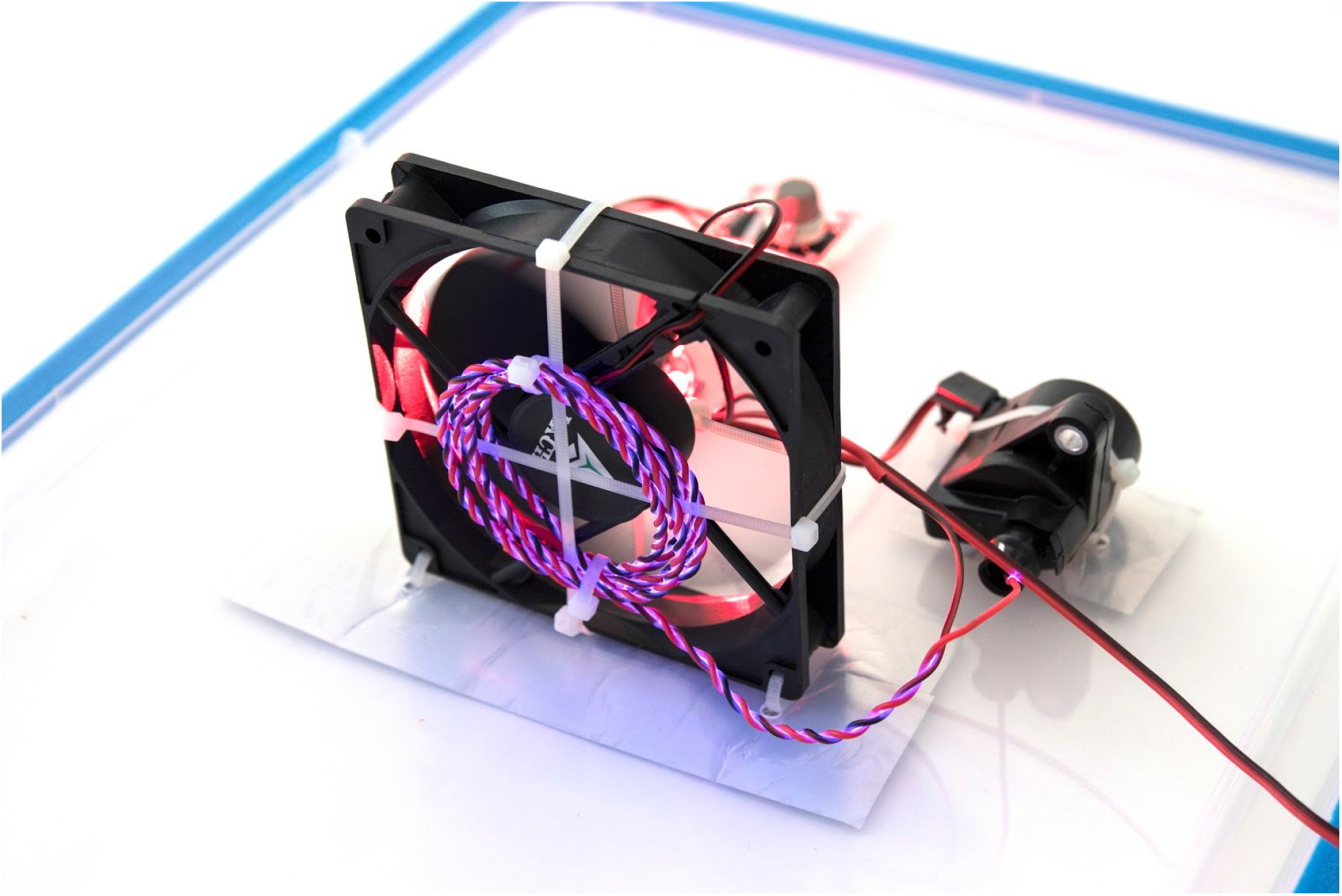
Long-exposure depiction of the ozone generation coil in service (corona discharge visible).

After all electronics are mounted to the inside of the container lid using zip ties, cover over all exposed holes in the chamber lid with aluminum HVAC tape in order to prevent ozone leaks from the chamber. The overall system should appear as shown in Fig. 9 (in this case ports for gas sampling are present, but this is not required). Optionally, a cover plate with labelling or instructions may also be added to the top of the chamber, as in the rendering of Fig. 10. Prior to treatment of items in a new ozone sterilization system, a burn-in period of three consecutive 150-minute cycles is recommended in order to ensure that the MQ-131 ozone sensor does not drift unacceptably over time. This step only needs to be performed once at the time of initial construction of the ozone sterilization system.

**Fig. 9.**
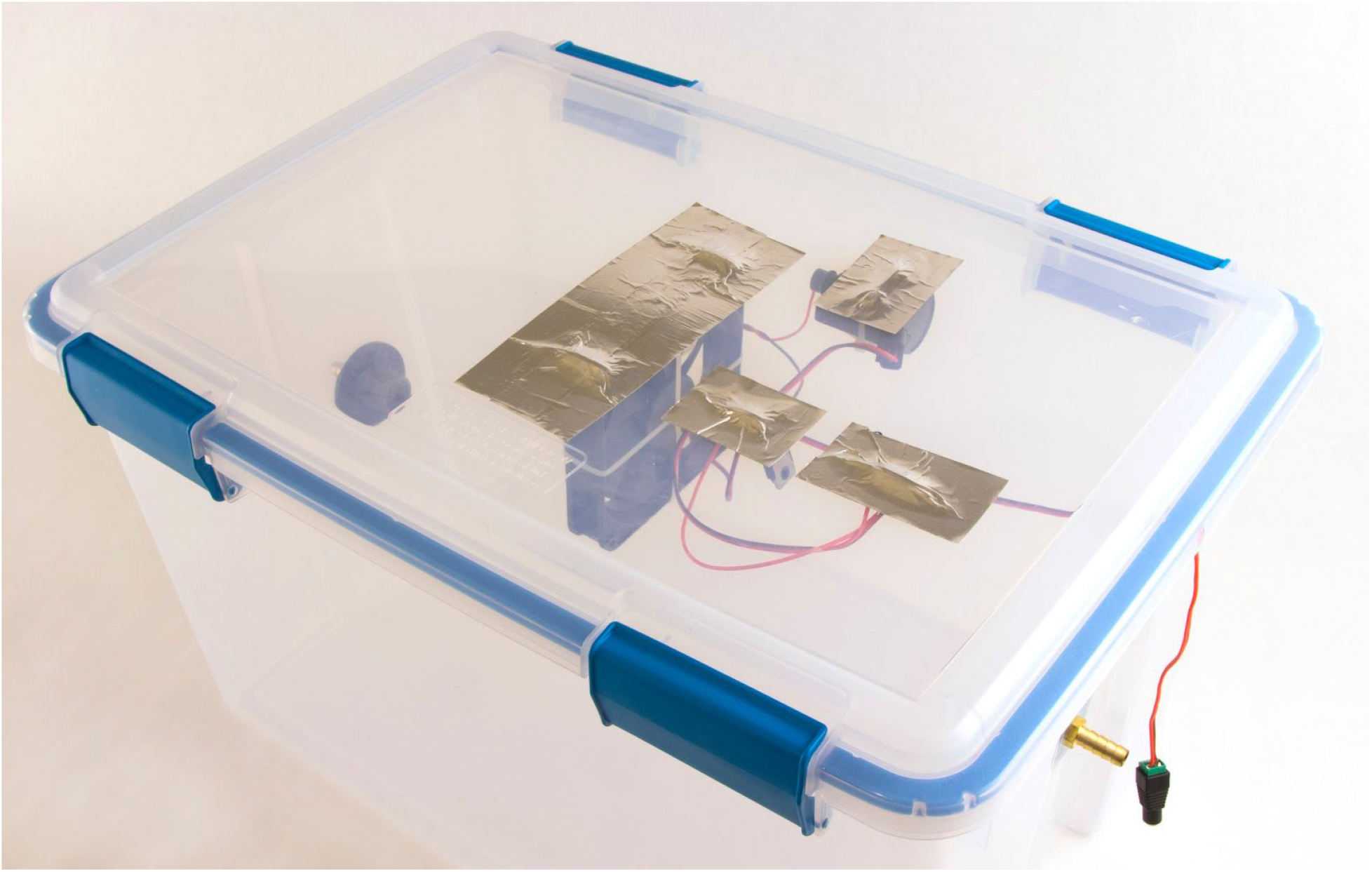
Outside of a complete closed-loop ozone gas generation system showing aluminum HVAC tap as sealing agent, no labelling attached.

**Fig. 10.**
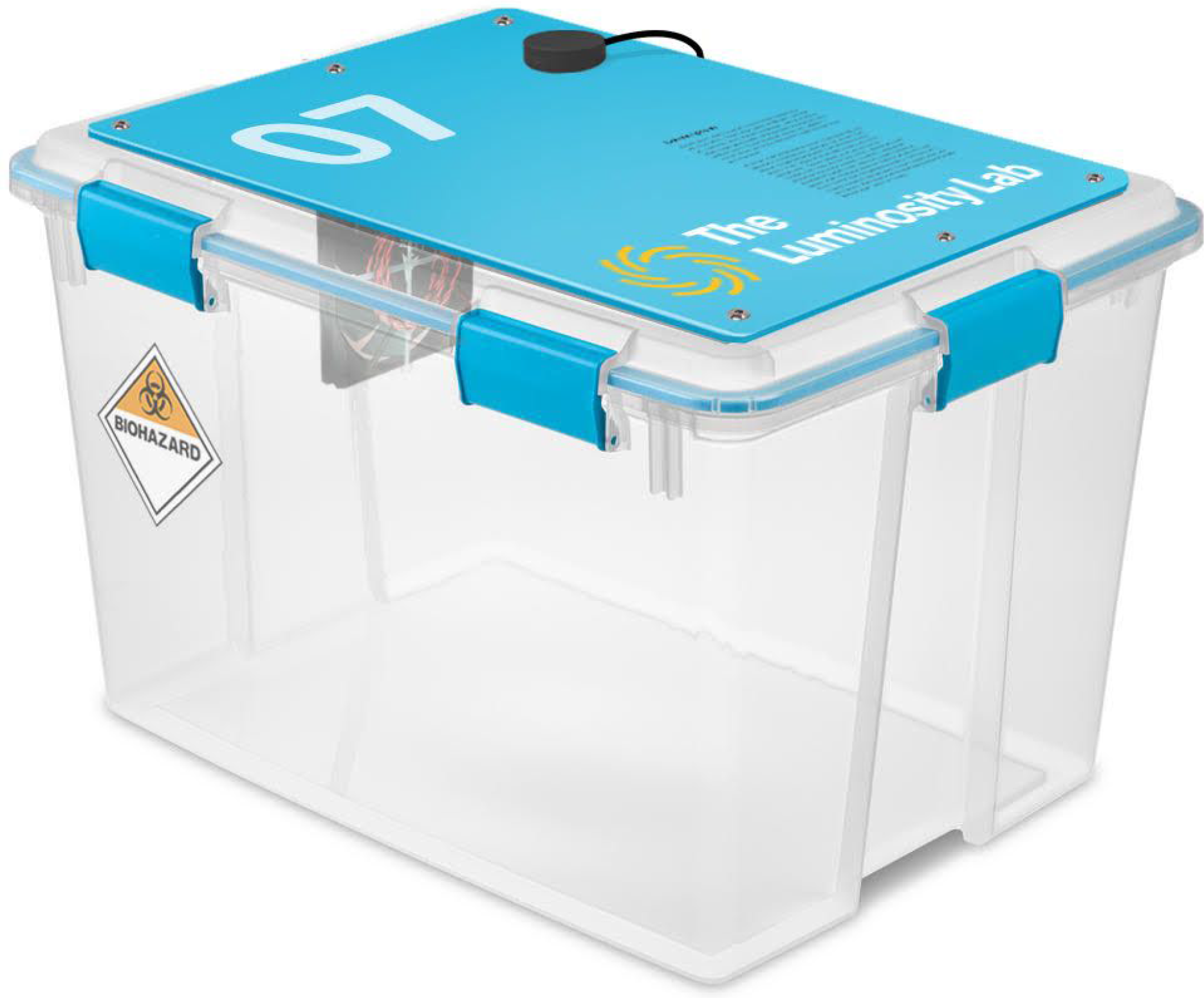
Digital rendering of a complete ozone gas sterilization system chamber with labelling and cover plate attached.

### Theory of circuit operation

The circuit depicted in Fig. 7 is designed to provide pulsed high voltage to the ozone generation coil using a flyback-type automotive ignition coil and NPN transistor. The NPN transistor is turned on and off by the microcontroller, typically at 980 Hz, using a pulse-width modulation (PWM) signal. When the transistor is turned on, electric current starts flowing in the primary winding of the spark ignition coil. The inductance of the spark ignition coil is high because it has an air gap, so the current through the coil rises gradually when voltage is applied to it by the NPN transistor turning on. When the NPN transistor turns off, the current through the spark ignition coil persists, resulting in a high voltage (∼190 V initially and 130V for a sustained period for the TIP35CW) being generated across the primary winding of the spark ignition coil. An oscilloscope plot of the voltage across the TIP35CW transistor during one cycle of operation is depicted in Fig. 11. Voltage is measured through a 10x attenuation probe, so voltage values are 10x the values shown here. The NPN transistor briefly operates in dielectric breakdown, which limits the peak voltage across the primary winding of the spark ignition coil. The limitation of voltage prevents damage to the coil, and also allows the flow of current to persist for longer, leading to a more sustained corona discharge at the ozone generation coil. This voltage is magnified by approximately 100x at the secondary winding of the transformer, which allows approximately 15 kV to be developed in the ozone generation coil. Corona discharge occurs and ozone is formed by the ionization and recombination of atmospheric oxygen.

**Fig. 11.**
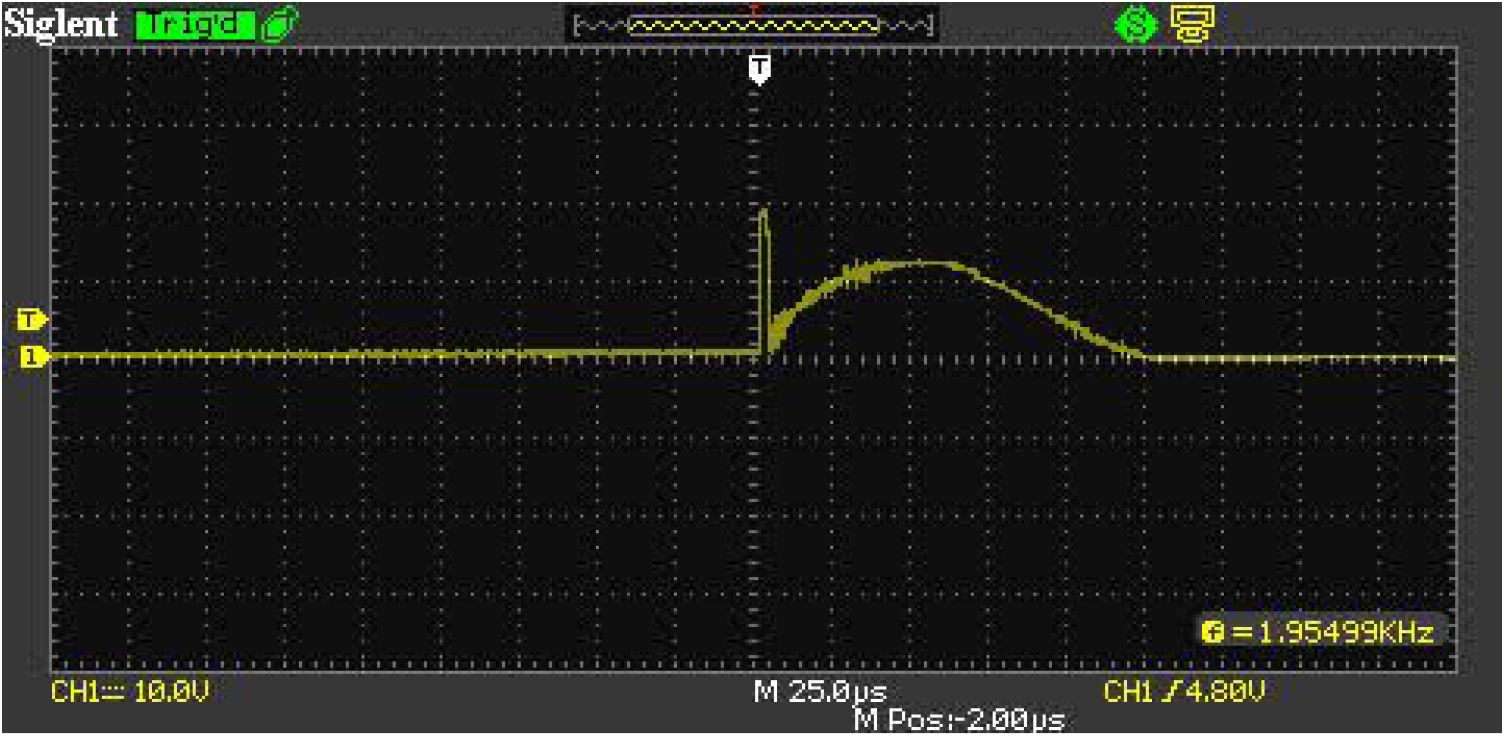
Oscilloscope plot showing the voltage between the collector and emitter of the TIP35CW during one cycle of operation. Vertical divisions are 100 V/div and horizontal divisions are 25 us/div.

### Microcontroller code

The C++ microcontroller code shown in Fig. 12 may be used in the Arduino IDE to program the Arduino Nano microcontroller to perform closed-loop control of the ozone concentration within the chamber at approximately 25 ppm. A suggested ADC value that has been verified with an MQ-131 sensor is provided, but for higher-precision applications this value may need to be trimmed using an ozone sensing system such as a *Thermo 49C* ozone analyzer at the time of unit commissioning.

**Fig. 12.**
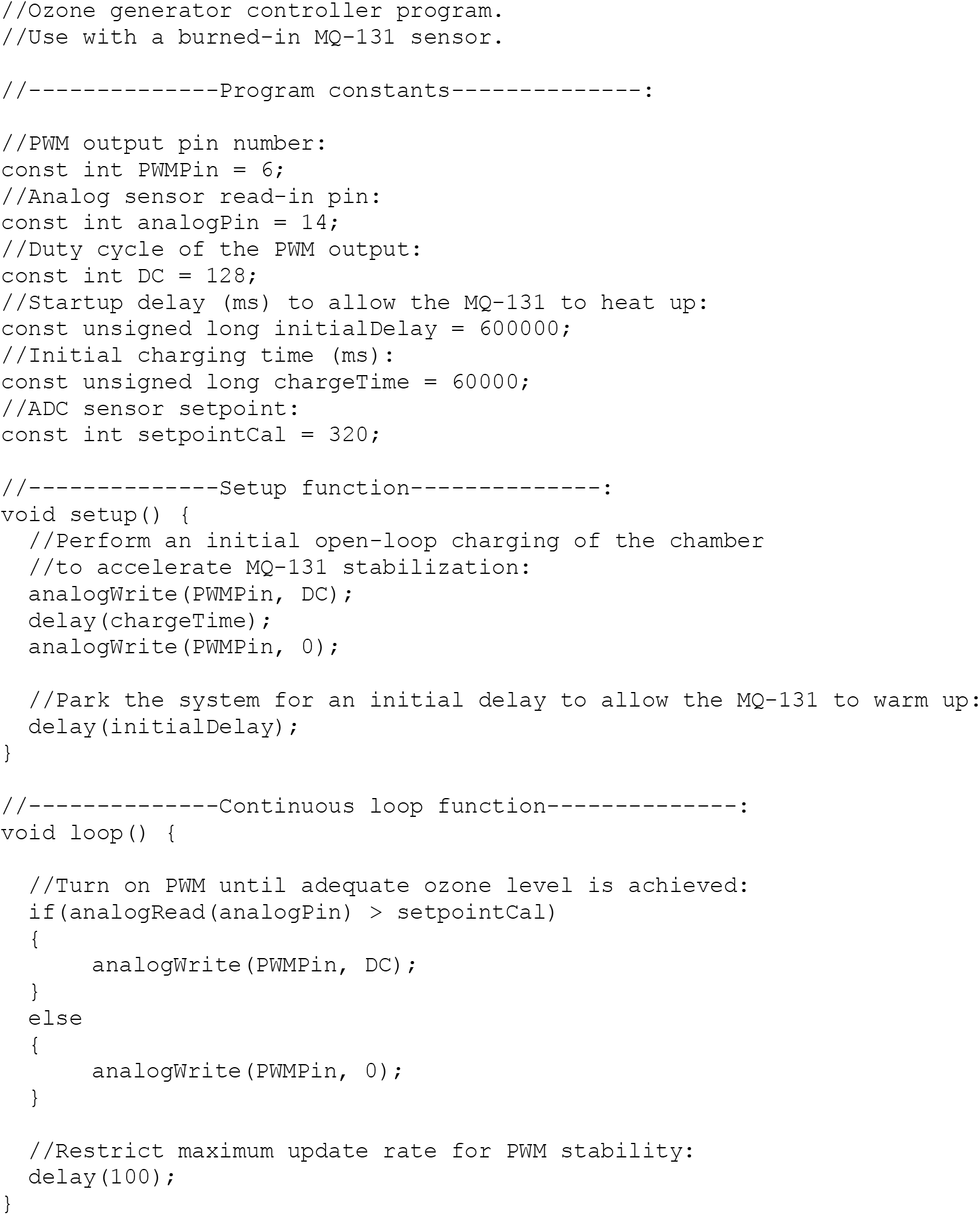
Arduino program code for operating a closed-loop implementation of the ozone treatment system.

The program is configured to initially charge the container with a small amount of ozone and then to allow the MQ-131 sensor to warm up for 10 minutes prior to closed-loop control.

This program works by generating a PWM signal at 50% duty cycle, which is sent to the TIP35CW transistor in order to facilitate the production of high voltage electricity at the terminals of the automotive ignition coil. The PWM signal is periodically turned on and off in order to maintain the ozone concentration within the chamber at the level specified by the analog sensor voltage setpoint, *VThresh*.

### Dosing for Sterilization

Table 4 outlines recommended dosages to achieve certain log reductions in viral contaminants, based on *Inactivation of Influenza Virus by Ozone Gas*, as most other viruses, including bacteriophages, are more sensitive to ozone than influenza [9]. Results during validation virucidal activity using P22 bacteriophages as a proxy for SARS-CoV-2 achieved a 6-log reduction and supported these recommended dosages.

### Considerations for the Safe Reuse of N95 Respirators

Certain precautions should be taken when reusing sterilized N95 respirators. This section should act only as a discussion of reusing ozone-treated N95s in the future if problems with elastic degradation are solved and further validation of sterilizing N95s without a significant effect on their performance is completed. The CDC recommends reuse of disposable N95s only during times of shortage [25]. With extended use of N95 respirators during epidemic-related shortages, respirators can experience significant wear and outside debris contamination that limit N95 performance and fit and reduce the ability of an N95 to be reused safely. With any sterilization and reuse of N95s, significant attention should be given to donning and doffing procedures to prevent contamination of healthcare personnel. A system for returning the respirators back to the original health care workers should be put in place, as outlined in reference 25. It is highly suggested to track the N95s by marking their containers to ensure each respirator has a single user, which reduces concerns about fit of the mask or complications such as the original user wearing cosmetics or lotion [26]. While a user seal check is required in many healthcare and industrial facilities, it is highly recommended that all personnel using a sterilized respirator should conduct one. At any visible deformation of shape, debris, or contaminate on the N95, the respirator should be disposed of [26].

The effect of VHP on the fluid resistance of the surgical N95 should only be a concern for healthcare professionals working in environments either requiring high sterility or with high risk of projectile bodily fluids, as the CDC suggests reserving surgical respirators for these instances during a shortage. Since N95s are only recommended to be reused in times of shortage, healthcare personnel not working in these settings would only require a non-surgical N95, which does not necessitate fluid resistance [27].

### System Operation

After a new system has been constructed and the described burn-in process has been performed, the system may be used to process items. Before treating large numbers of identical items or respirators, it is recommended to run an initial test cycle with one unit of the specific item to be sterilized to ensure compatibility.

To begin treatment, place materials into the system. Ensure clearance on all sides of items in the chamber. Next, secure the lid on the top of the chamber using the provided clasps. Plug in the ozone generator module. The ozone generator should generate a high-pitched whine for several seconds, indicating a successful start-up. The system will cycle and off throughout the treatment period. Treatment times are given as shown in Table 4. When the treatment period has concluded, unplug the system from the power source. Keeping your face turned away from the system, open the lid in a well-ventilated space. Allow approximately 10 minutes for the ozone to dissipate, then remove the items being decontaminated. It is important to allow respirators or any face worn coverings to aerate before being donned by personnel.

